# Comprehensive Analysis of Atypical chronic myeloid leukemia (aCML): Epidemiology, Clinical Features, and Survival Outcomes Based on SEER Database Insights

**DOI:** 10.1101/2024.07.28.24311130

**Authors:** Fan Wang

## Abstract

**Background:** Atypical Chronic Myeloid Leukemia (aCML) is a rare and aggressive myelodysplastic syndrome/myeloproliferative neoplasm. This study aimed to provide a comprehensive understanding of the epidemiology, clinical characteristics, and survival outcomes of aCML patients.

**Methods:** The study utilized data from the Surveillance, Epidemiology, and End Results (SEER) database from 2001 to 2020. The age-adjusted incidence rate (AIR) of aCML was calculated, and survival outcomes were analyzed using the Kaplan-Meier method and accelerated failure time (AFT) regression analysis.

**Results:** The AIR of aCML was found to be 0.024 per 100,000 person-years, with the highest rate observed in 2020. The incidence of aCML increased with age and was higher in males. The study cohort predominantly consisted of elderly White individuals, with an average age at diagnosis of 68.2 ± 15.3 years. The median overall survival (OS) and disease-specific survival (DSS) were 1.4 years and 1.7 years, respectively. Older age was independently associated with worse survival outcomes. Notably, treatment delay and chemotherapy did not significantly impact OS or DSS.

**Conclusions:** This study provides comprehensive insights into the epidemiology, clinical characteristics, and survival outcomes of aCML, highlighting its rarity, aggressive nature, and poor prognosis. Further research is needed to validate these findings and explore novel therapeutic strategies for improving outcomes in this challenging hematologic malignancy.

## Introduction

Atypical chronic myeloid leukemia (aCML) is a rare and aggressive hematologic malignancy that belongs to the category of myelodysplastic/myeloproliferative (MDS/MPN) neoplasms, which affect the bone marrow and the blood cells, causing both abnormal proliferation and impaired maturation of blood cells^1,2^. It is characterized by leukocytosis with dysgranulopoiesis, anemia and/or thrombocytopenia, splenomegaly^3^. According to the WHO diagnostic criteria for aCML, the white blood count is ≥13 × 10^9^/L, with immature granulocytes ≥10% of leukocytes and < 20% blasts in the blood and bone marrow^1,2^. Unlike the more common chronic myeloid leukemia (CML), aCML does not possess the Philadelphia chromosome or the BCR-ABL1 fusion gene, which is the hallmark genetic abnormalities that drive CML^4^. aCML also does not have other known gene rearrangements that can cause similar diseases, such as PDGFRA, PDGFRB, FGFR1 or PCM1-JAK2^5^. In aCML, the so-called MPN-associated driver mutations (JAK2, CALR, MPL) are generally not present as well^1^. However, aCML can be associated with other cytogenetic changes including oncogenic mutations in the granulocyte-colony stimulating factor receptor (CSF3R)^4^ and isochromosome 17q^6^. Moreover, in up to 1/3 of cases, aCML is linked to mutations in SETBP1 or ETNK1^7,8^.

The diagnosis of aCML is challenging due to its overlapping features with other MDS/MPN entities, such as chronic myelomonocytic leukemia (CMML), atypical chronic myeloid leukemia (CNL), and MDS/MPN not otherwise specified (MDS/MPN NOS)^2,9,10^. The diagnosis of aCML is based on blood and bone marrow tests that show less than 20% blasts in the peripheral blood and bone marrow and the presence of leukocytosis, dysgranulopoiesis, and the absence of BCR-ABL1 or other gene rearrangements that can cause similar diseases^11^. The prognosis of aCML is generally poor, with a median overall survival of less than 2 years after diagnosis^12^. The disease tends to progress rapidly, with a high likelihood of transformation into acute myeloid leukemia (AML)^13,14^. The traditional prognostic scoring systems, such as the International Prognostic Scoring System (IPSS) used in myelodysplastic syndromes, are not fully applicable to aCML due to its unique clinical and genetic features^10^. The main factors that affect the prognosis are the age of the patient, the percentage of blasts in the blood or bone marrow, and the presence of certain genetic mutations, such as ASXL1, SETBP1, or RUNX1^15,16^. The prognosis of aCML has not improved over the last 20 years, partly because of the lack of specific recurrent genomic or karyotypic abnormalities that can be targeted by therapy^14^. The molecular pathogenesis of aCML remains elusive and heterogeneous, involving multiple pathways and genes that regulate cell growth, differentiation, and survival^16^. Therefore, there is an urgent need for more research and clinical trials to identify novel biomarkers and therapeutic targets for aCML.

The treatment of aCML is challenging because there is no established standard of care or effective therapy for this disease^11^. The main goal of treatment is to control the symptoms, prevent complications, and improve the quality of life of the patients^11^. Allogeneic hematopoietic stem cell transplantation (HSCT) is the only potentially curative option for eligible patients with a suitable donor^16^. However, the optimal timing and criteria for HSCT are not well defined^16^. Non-transplant therapies for aCML have been largely empirical, based on limited data from case series or retrospective studies^16^. These include conventional cytotoxic agents, hypomethylating agents, interferon-alpha, and tyrosine kinase inhibitors^16^. In recent years, advances in molecular characterization of aCML have revealed recurrent mutations in genes involved in epigenetic regulation, signal transduction, transcriptional regulation, and splicing^17^. These mutations may have diagnostic, prognostic, and therapeutic implications for aCML patients^18^. Continued research is crucial to better understand aCML’s biology and develop more effective, targeted therapies.

However, aCML remains poorly understood due to its rarity. The purpose of this study is to explore the epidemiological characteristics and identify the factors affecting the survival of aCML patients based on a population-based study using the national Cancer Institute’s Surveillance, Epidemiology and End Results (SEER) database.

## Materials and Methods

### Data collection for aCML study

Data for this study were collected from the Surveillance, Epidemiology, and End Results (SEER) Program (https://seer.cancer.gov/) maintained by the National Cancer Institute (NCI)^19^. The SEER*Stat software version 8.4.3 (https://seer.cancer.gov/seerstat/, accessed on June 10, 2024) was used to retrieve the data^20^. The SEER 17 database [Incidence-SEER Research Data, 17 registries, Nov 2022 Sub (2000-2020)] was utilized to extract data of disease incidence by using the “rate session”. Through the “Incidence-SEER Research Plus Data, 17 Registries, Nov 2022 Sub (2000-2020),” which covers approximately 26.5% of the U.S. population (based on 2020 census), patients diagnosed with aCML between 2001 and 2020 were selected using the case listing session, and cases with only known age (censored at age 89 years) and only malignant behavior were taken in. As shown in the flow chart (Figure 1), the inclusion criteria of aCML patients were as follows: (1) the International Classification of Diseases for Oncology (ICD-O-3) histologic code (9876/3); (2) the patient’s survival time was not 0 or unknown. The exclusion criteria were as follows: (1) the type of reporting source was “laboratory only”; (2) the diagnosis confirmation was “clinical diagnosis only” or unknown. A total of 255 patients with aCML were finally included in the final cohort. The ethical approval from the ethics committee was not required as the SEER database are anonymized and publicly available.

**Figure 1.**
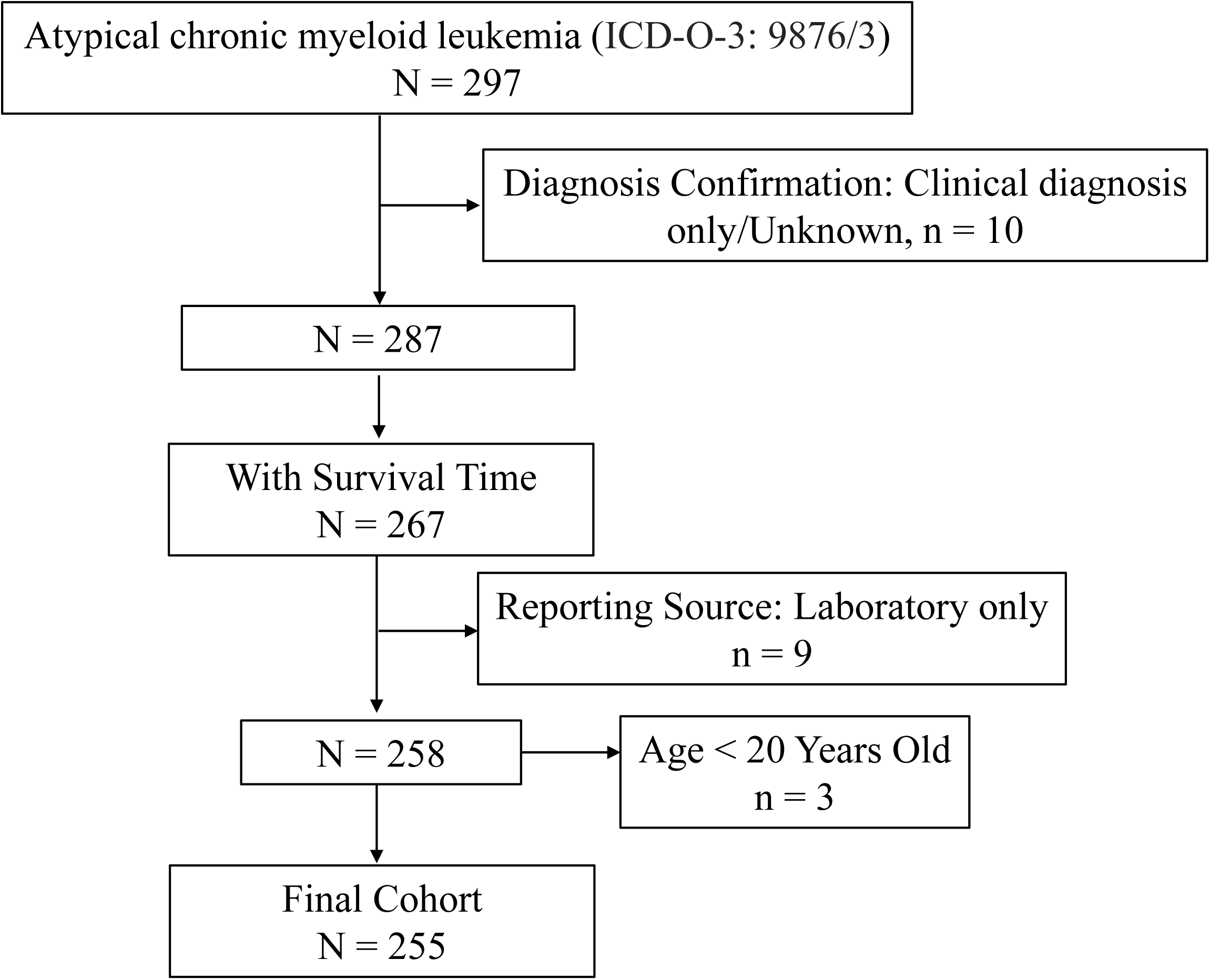
Flow chart of study cohort selection using the SEER database. A flow diagram of aCML patient selection in this study. aCML, atypical chronic myeloid leukemia; SEER, Surveillance, Epidemiology, and End Results.

### Variable Definition of aCML

The variables collected for analysis included age, sex, race, marital status, year of diagnosis, primary site, vital status, survival months, cause-specific death classification, cause of death to site, sequence number, first malignant primary indicator, total number of in situ/malignant tumors for patient, type of reporting source, diagnostic confirmation, chemotherapy recode, and so on. Age was categorized as < 60 years old or 60+ years old at aCML diagnosis. Race was classified as African American, White, or Other (Asian/Pacific Islander and American Indian/Alaska Native). Marital status included categories of married, single, and other (comprising Divorced, Separated, and Widowed). Income was taken from the variable “Median Household Income in the past 12 months (in 2021 Inflation Adjusted Dollars)” and categorized as <$50,000, $50,000-$75,000, and $75,000+. According to the size of the population and the level of economic development, residence type was divided into two categories: living in a metropolitan area with a population of 1 million or more and other including living in a metropolitan area with a population of about 250,000 to 1 million, live in a metropolitan area with a population of less than 250,000, living in non-metropolitan areas. The variable “sequence number” was classified into two groups: primary aCML and secondary aCML (arises due to other primary malignancies). Cause-of-death information was extracted from the “COD to site recode” field. In the SEER database, aCML-related deaths were classified as those attributed directly to the cancer diagnosis (“dead (attributed to this cancer diagnosis)”). Conversely, aCML-unrelated deaths were categorized as deaths resulting from causes other than the cancer diagnosis (“dead (attributable to causes other than this cancer diagnosis)”). Overall survival time was calculated from the date of aCML diagnosis until death or last follow up.

### Statistical analysis

Statistical analyses were conducted using R version 4.2.1 (http://www.r-project.org/, R Foundation for Statistical Computing, Vienna, Austria). Baseline characteristics of aCML patients were compared between groups defined as “aCML-related death” and “other cause”. Continuous data were analyzed using Student’s *t*-test, while categorical variables were assessed with Chi-square tests; Fisher’s exact test was utilized when cell frequencies were insufficient. Overall survival (OS) and disease-specific survival (DSS) were estimated using Kaplan-Meier methods with log-rank tests. Due to violations of the Cox proportional hazards regression assumption by Schoenfeld residual test, parametric univariate and multivariable accelerated failure time (AFT) regression analyses with the Weibull distribution were performed. All variables that underwent AFT univariate analysis were included in the multivariable AFT model to account for any confounding effects. The resulting coefficients from the AFT analysis were converted into hazard ratios (HR) and 95% confidence intervals (95% CIs) for easier interpretation. All *P* values were two-sided and a *P* value < 0.05 was defined as statistically significant.

## Results

### Incidence of aCML

Analysis of the SEER database indicated that the age-adjusted incidence rate (AIR) of aCML from 2000 to 2020 [age adjusted to the 2000 US Standard Population (19 age groups - Census P25-1130)] was 0.024 per 100,000 person-years (95% CI, 0.022-0.027). The annual variation in AIR is depicted in Figure 2A, showing the highest rate in 2020 at 0.038 per 100,000 person-years (95% CI, 0.025-0.055). Notably, the incidence of aCML increased with age, as illustrated in Figure 2B. Compared to patients under 60 years old (AIR 0.006/100,000 person-years), the incidence rate ratio (IRR) for those aged 60 and older (AIR 0.084/100,000 person-years) was 18.23 (*P* < 0.0001). Regarding gender differences, males exhibited a significantly higher AIR (0.035/100,000 person-years) compared to females (AIR 0.017/100,000 person-years, IRR 2.09, 95% CI, 1.64-2.68, *P* < 0.0001; Figure 2C, D).

**Figure 2.**
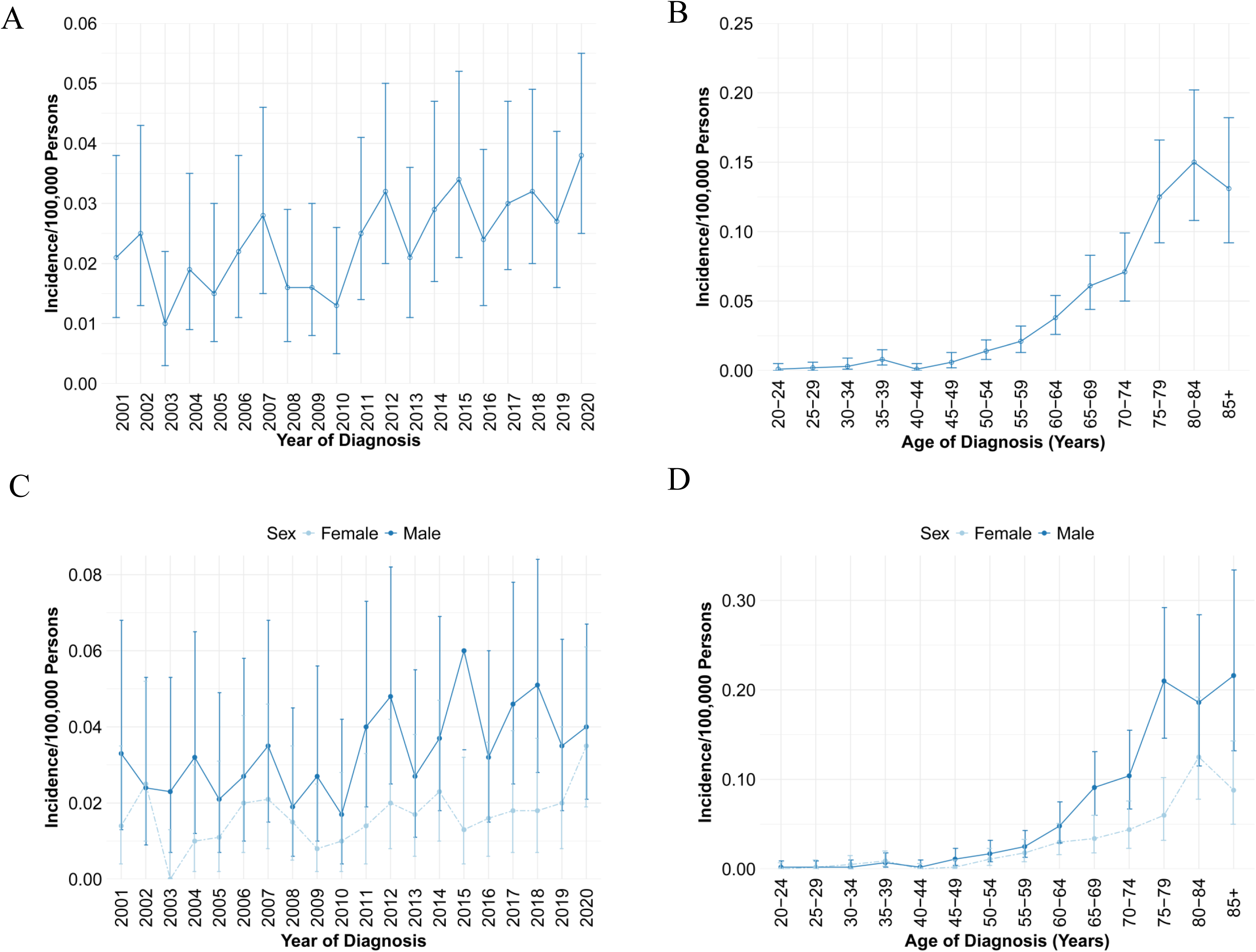
Age-adjusted incidence of aCML from 2001 to 2020 in SEER database. (A) Annual age-adjusted incidence of aCML; (B) Age-adjusted incidence of aCML based on age of diagnosis; (C) Annual age-adjusted incidence of aCML in male and female populations, respectively; (D) Age-adjusted incidence of aCML based on age of diagnosis in male and female patients, respectively. aCML, atypical chronic myeloid leukemia.

### Demographics of aCML Patients

As shown in Figure 1, a total of 255 patients were finally identified as aCML in the SEER 17 registry, Nov 2022 Sub (2000-2020) from January 2001 to December 2020. The primary site of manifestation was the bone marrow for all patients (*n* = 255, 100%). For all patients in the study cohort, 62.4% were male, about 1.7 folds that of female (*n*=96, 37.6%; Table 1). The average age at diagnosis was approximately 68.2 ± 15.3 years, with 22.0% aged less than 60 years (< 60) and 78.0% aged 60 or older (60+). The majority of aCML patients were White (80.0%), while African American and Other racial groups (including Asian/Pacific Islander and American Indian/Alaska Native) comprised 9.0% and 11.0%, respectively. At diagnosis, 57.3% of patients were married, 31.8% had other marital statuses (divorced, separated, or widowed), and 11.0% were single and had never married. The highest incidence spanned the years 2011-2020 (65.9%). Of all cases, 74.5% were classified as primary aCML, while 25.5% were secondary to other primary malignancies. Regarding income distribution, 43.9% of patients reported a median household income exceeding $75,000, 44.7% reported incomes between $50,000 and $75,000, and 11.4% reported incomes below $50,000. The majority of patients resided in metropolitan areas with populations of 1 million or more (54.9%). Approximately 80.8% of aCML patients received chemotherapy. At the last follow-up, 57 (22.4%) patients were alive; 159 (62.4%) deaths were attributed to aCML, while an additional 39 (15.3%) patients died from other causes such as diseases of heart (n=12), cerebrovascular diseases (n=1), accidents and adverse effects (n=3), and so on. Details on the distribution of causes of death unrelated to aCML are summarized in Table 1. Importantly, no significant differences were found between the “aCML-related death” and “other cause” survival groups. The epidemiological characteristics and survival comparisons of aCML are presented in Table 2.

### Survival Analysis of aCML

During the follow-up period, there were 198 deaths, with 159 deaths attributed to disease-specific causes. As shown in Figure 3A and B, the median survival time for OS and DSS was 1.4 years and 1.7 years, respectively; the 1-year, 3-year, 5-year, and 10-year OS rates were 62.4%, 24.7%, 17.8%, and 11.2%, respectively, and the corresponding DSS rates were 67.3%, 31.1%, 25.7%, and 18.9%, respectively. While patients diagnosed in 2011-2020 appeared to have worse OS than those diagnosed in 2001-2010, further analysis revealed no significant differences between the diagnosis years (*P* = 0.400; Figure 3C). Similarly, there were neither significant difference for DSS between the diagnosis years (*P* = 0.660; Figure 3D). Furthermore, Kaplan-Meier analysis of OS indicated that only older age was associated with poorer overall survival (*P* < 0.0001; Figure 4A). No significant differences in OS were observed by sex (*P* = 0.960, Figure 4B), race (*P* = 0.340, Figure 4C), marital status (*P* = 0.057, Figure 4D), sequence (*P* = 0.420, Figure 4E), income (*P* = 0.880, Figure 4F), residence (*P* = 0.380, Figure 4G), chemotherapy (*P* = 1.000, Figure 4H) or treatment delay (*P* = 0.890, Figure 4I). Similarly, Kaplan-Meier analysis of DSS showed consistent results (Figure 5).

**Figure 3.**
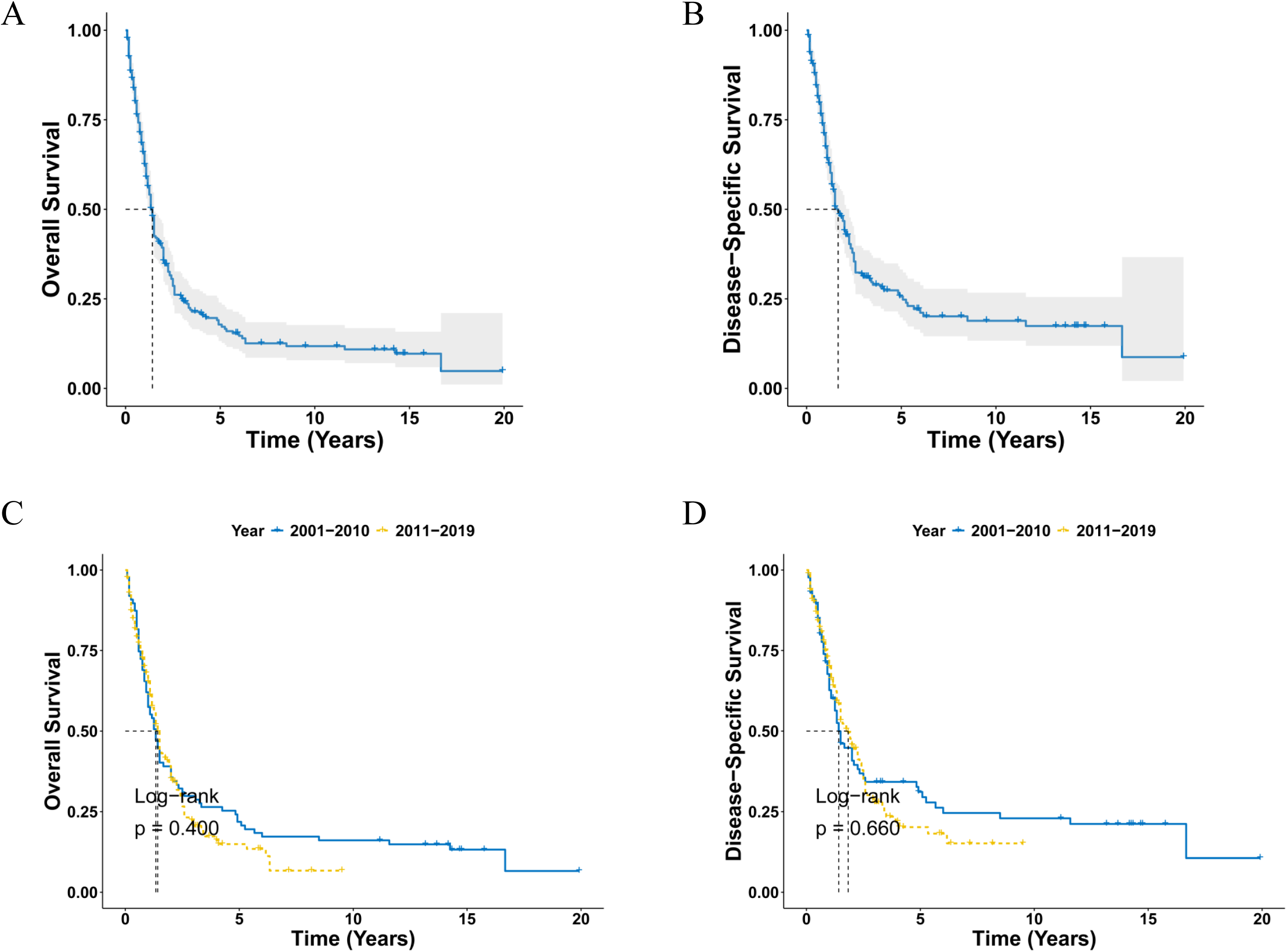
Survival analysis of aCML. **(A, B)** OS (A) and DSS (B) curves for all aCML patients. **(C, D)** Survival curves of OS (C) and DSS (D) according to the diagnosis years. aCML, atypical chronic myeloid leukemia; OS, overall survival; DSS, disease-specific survival.

**Figure 4.**
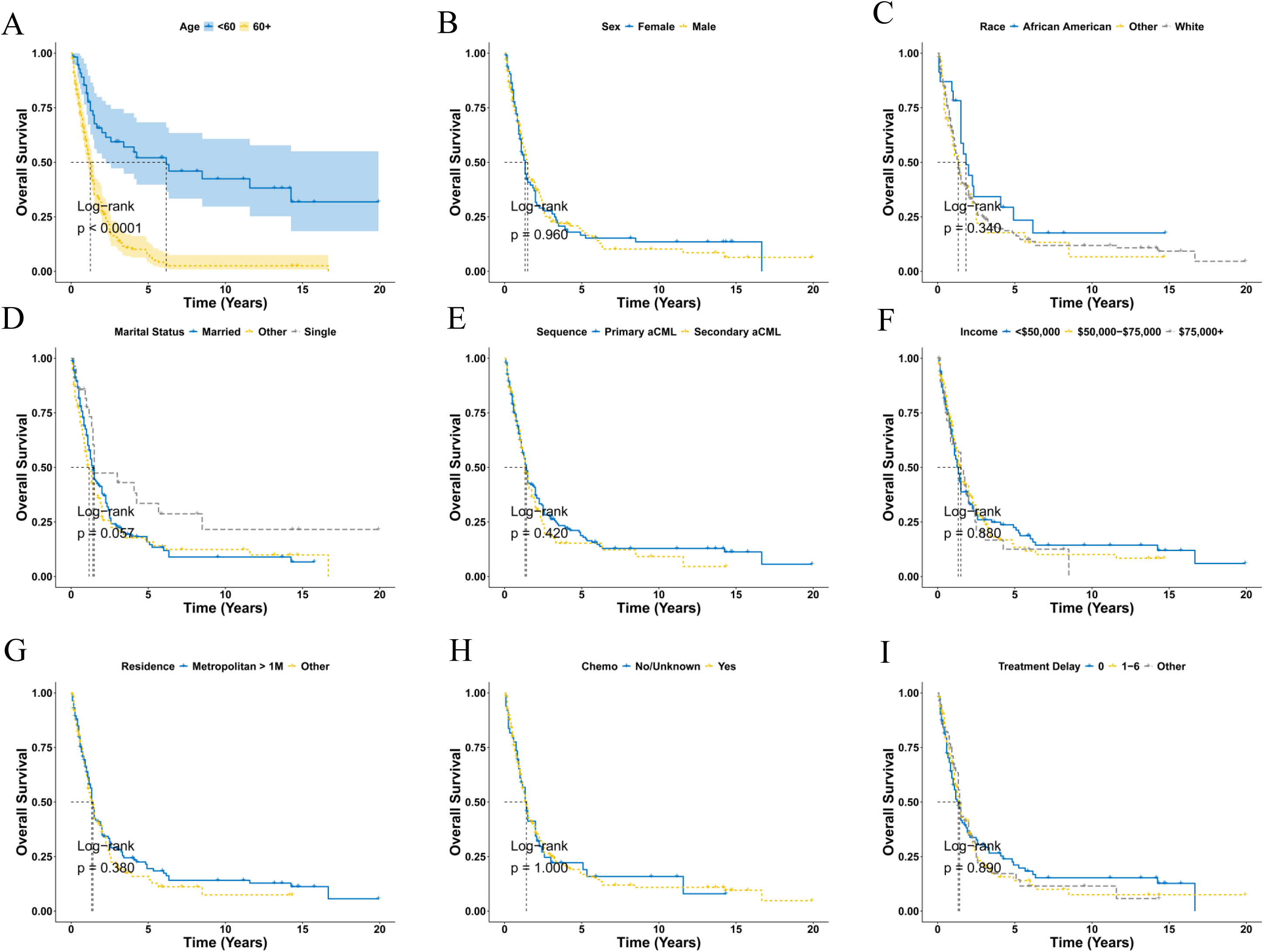
Overall survival analysis of aCML stratified by age (A), sex (B), race (C), marital status (D), sequence (E), income (F), residence (G), chemotherapy (H) and treatment delay (I) using Kaplan-Meier method. aCML, atypical chronic myeloid leukemia.

**Figure 5.**
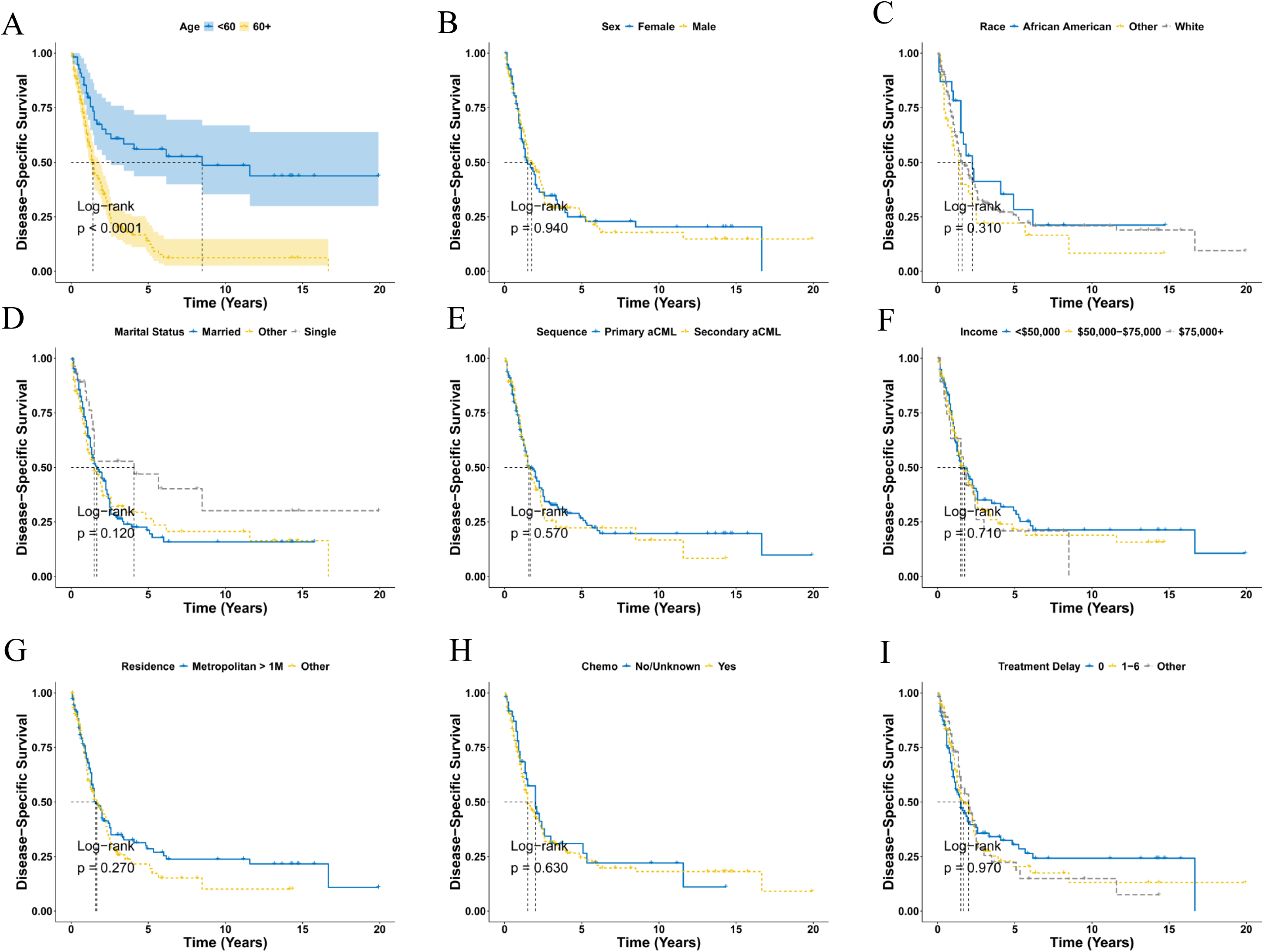
Disease-specific survival analysis of aCML stratified by age (A), sex (B), race (C), marital status (D), sequence (E), income (F), residence (G), chemotherapy (H) and treatment delay (I) using Kaplan-Meier method. aCML, atypical chronic myeloid leukemia.

### Univariate and Multivariable AFT Regression Analysis of aCML

The univariate AFT regression analysis of OS showed only older age was significantly associated with worse outcomes (Table 3). Other factors such as sex, race, marital status diagnosis year, sequence, income, residence, treatment delay and chemotherapy did not impact the OS. Similar results were obtained with the univariate AFT regression analysis of DSS (Table 3). In the multivariable AFT regression model including all univariate variables (Table 4), older age remained independently associated with worse OS (HR: 3.65, CI=2.34-5.70, *P* < 0.001). Similar findings were observed for DSS (HR: 3.08, CI=1.92-4.90, *P* < 0.001; Table 4).

## Discussion

In conclusion, this study presents a comprehensive analysis of the incidence, demographics, and survival outcomes of atypical chronic myeloid leukemia (aCML) using data from the SEER database. The observed age-adjusted incidence rate (AIR) of aCML from 2000 to 2020 aligns with previous reports, reaffirming its rarity within the spectrum of myelodysplastic syndrome (MDS)/myeloproliferative neoplasm (MPN)^11^. Our findings indicate a temporal variability in incidence, with the highest rates documented in 2020, underscoring the influence of evolving disease surveillance and diagnostic practices on reported trends. Importantly, the incidence of aCML increases with age and is more prevalent among males, consistent with existing literature highlighting age and gender as significant risk factors^21^.

Our study highlights a predominant occurrence of aCML among elderly individuals, with an average diagnosis age of 68.2 years, reflecting advanced age as a notable risk factor for disease onset and progression^22,23^. Additionally, the predominance of White patients and the distribution across diverse socioeconomic strata underscore the multifactorial nature of disease epidemiology, influenced by genetic, environmental, and healthcare access factors.

Survival analysis revealed challenging outcomes for aCML patients, with median overall survival (OS) and disease-specific survival (DSS) times of 1.4 years and 1.7 years, respectively, highlighting the aggressive nature of the disease. While temporal trends suggested worsened OS in patients diagnosed from 2011 to 2020 compared to earlier cohorts, statistical analysis did not identify significant differences, suggesting persistent challenges in improving outcomes over time^24^. Multivariate regression analyses confirmed older age as a significant independent predictor of poorer OS and DSS, consistent with prior studies emphasizing age as a critical determinant of disease trajectory^22,24^. Conversely, factors such as sex, race, marital status, and treatment modalities did not significantly influence survival outcomes in our cohort, aligning with literature emphasizing age-related factors as primary drivers of clinical outcomes^25^.

An intriguing finding of our study is the lack of significant impact of chemotherapy on OS or DSS among aCML patients, contrary to expectations based on treatment efficacy in other hematologic malignancies^26^. This discrepancy suggests potential intrinsic resistance mechanisms within aCML, such as genetic mutations or altered signaling pathways, which may diminish treatment effectiveness^27^. Furthermore, the advanced age and prevalent comorbidities among our patient population likely contributed to suboptimal treatment responses, emphasizing the need for personalized therapeutic approaches tailored to individual patient profiles^16^.

Another unexpected finding was the absence of a significant association between treatment delay and survival outcomes in aCML patients. This contrasts with general oncologic principles where delayed treatment initiation typically correlates with poorer prognosis^28,29^. Potential reasons for this observation include the rapid disease progression inherent to aCML, where the timing of treatment initiation may not substantially alter clinical outcomes. Moreover, limitations in the study’s ability to capture detailed information on treatment delays and their specific impacts on disease progression further complicate interpretation.

However, several limitations warrant consideration. The retrospective nature of our study using SEER database limits causal inference, and the lack of comprehensive clinical data, including molecular and cytogenetic profiles, precludes a deeper understanding of disease biology and its impact on outcomes. Moreover, the study’s reliance on registry data may introduce biases, and the exclusion of patients with incomplete records may affect the generalizability of our findings. Additionally, the study does not account for the influence of comorbidities on survival outcomes, which could confound interpretations.

## Conclusions

In conclusion, this study provides critical insights into the epidemiology, clinical characteristics, and treatment outcomes of aCML, highlighting persistent challenges in achieving favorable outcomes with standard therapies. Our findings underscore the urgent need for further research to validate these observations in diverse populations and settings. Future research efforts should prioritize elucidating molecular mechanisms of treatment resistance, exploring novel therapeutic targets, and conducting prospective studies to validate findings and guide the development of tailored treatment strategies for this rare and aggressive hematologic malignancy.

## Data Availability Statement

The data analyzed in this study are from the SEER database (https://seer.cancer.gov/) that are available to the public.

## Acknowledgments

The interpretation of the data is the sole responsibility of the author(s). The author(s) acknowledge the efforts of the National Cancer Institute and the Surveillance, Epidemiology, and End Results (SEER) Program tumor registries in the creation of the SEER database.

## Abbreviations

AFT, accelerated failure time; aCML, Atypical chronic myeloid leukemia; Chemo, chemotherapy; CI, confidence interval; COD, cause of death; DSS, disease-specific survival; HR, hazard ration; OS, overall survival; RT, radiation therapy; SEER, Surveillance, Epidemiology, and End Results; NOS, not otherwise specified.

