## Supplementary material for "Comprehensive Analysis of Atypical chronic myeloid leukemia (aCML): Epidemiology, Clinical Features, and Survival Outcomes Based on SEER Database Insights": Table 1

**Table 1** Distribution of the cause of aCML-unrelated deaths.

| **Cause of Death** | **All Patients (N=39)** |
| --- | --- |
| Accidents and Adverse Effects | 3 (7.7%) |
| Alzheimers (ICD-9 and 10 only) | 1 (2.6%) |
| Breast | 1 (2.6%) |
| Cerebrovascular Diseases | 1 (2.6%) |
| Chronic Liver Disease and Cirrhosis | 1 (2.6%) |
| Chronic Obstructive Pulmonary Disease and Allied Cond | 3 (7.7%) |
| Colon excluding Rectum | 2 (5.1%) |
| Diabetes Mellitus | 1 (2.6%) |
| Diseases of Heart | 12 (30.8%) |
| Intrahepatic Bile Duct | 1 (2.6%) |
| Liver | 1 (2.6%) |
| Lung and Bronchus | 1 (2.6%) |
| Other Cause of Death | 6 (15.4%) |
| Other Infectious and Parasitic Diseases including HIV | 1 (2.6%) |
| Pneumonia and Influenza | 1 (2.6%) |
| Prostate | 1 (2.6%) |
| Suicide and Self-Inflicted Injury | 1 (2.6%) |
| Symptoms, Signs and Ill-Defined Conditions | 1 (2.6%) |

aCML, atypical chronic myeloid leukemia.
