## Supplementary material for "Comprehensive Analysis of Atypical chronic myeloid leukemia (aCML): Epidemiology, Clinical Features, and Survival Outcomes Based on SEER Database Insights": Table 2

Table 2 Baseline characteristics of patients with aCML.

| **Characteristics** | **Total (N=255)** | **Alive (N=57)** | **Death** | | ***P*-value** |
| --- | --- | --- | --- | --- | --- |
|  |  |  | **aCML-related Death**^10^ **(N=159)** | **Other cause**^11^ **(N=39)** |  |
| **Sex*, n (%)*** |  |  |  |  |  |
| Male | 159 (62.4%) | 34 (59.6%) | 100 (62.9%) | 25 (64.1%) | 1.00 |
| Female | 96 (37.6%) | 23 (40.4%) | 59 (37.1%) | 14 (35.9%) |  |
| **Age*, n (%)*** |  |  |  |  |  |
| <60 | 56 (22.0%) | 27 (47.4%) | 25 (15.7%) | 4 (10.3%) | 0.54 |
| 60+ | 199 (78.0%) | 30 (52.6%) | 134 (84.3%) | 35 (89.7%) |  |
| **Race*, n (%)*** |  |  |  |  |  |
| White | 204 (80.0%) | 45 (78.9%) | 123 (77.4%) | 36 (92.3%) | 0.10 |
| African American | 23 (9.0%) | 6 (10.5%) | 15 (9.4%) | 2 (5.1%) |  |
| Other^1^ | 28 (11.0%) | 6 (10.5%) | 21 (13.2%) | 1 (2.6%) |  |
| **Diagnosis Year*, n (%)*** |  |  |  |  |  |
| 2001-2010 | 87 (34.1%) | 11 (19.3%) | 62 (39.0%) | 14 (35.9%) | 0.86 |
| 2011-2020 | 168 (65.9%) | 46 (80.7%) | 97 (61.0%) | 25 (64.1%) |  |
| **Sequence*, n (%)*** |  |  |  |  |  |
| Primary aCML^2^ | 190 (74.5%) | 44 (77.2%) | 118 (74.2%) | 28 (71.8%) | 0.92 |
| Secondary aCML^3^ | 65 (25.5%) | 13 (22.8%) | 41 (25.8%) | 11 (28.2%) |  |
| **Marital Status*, n (%)*** |  |  |  |  |  |
| Married | 146 (57.3%) | 33 (57.9%) | 94 (59.1%) | 19 (48.7%) | 0.45 |
| Single^4^ | 28 (11.0%) | 10 (17.5%) | 14 (8.8%) | 4 (10.3%) |  |
| Other^5^ | 81 (31.8%) | 14 (24.6%) | 51 (32.1%) | 16 (41.0%) |  |
| **Income**^6^***, n (%)*** |  |  |  |  |  |
| <$50,000 | 29 (11.4%) | 5 (8.8%) | 20 (12.6%) | 4 (10.3%) | 0.57 |
| $50,000-$75,000 | 114 (44.7%) | 23 (40.4%) | 70 (44.0%) | 21 (53.8%) |  |
| $75,000+ | 112 (43.9%) | 29 (50.9%) | 69 (43.4%) | 14 (35.9%) |  |
| **Residence*, n (%)*** |  |  |  |  |  |
| Metropolitan > 1M^7^ | 140 (54.9%) | 31 (54.4%) | 85 (53.5%) | 24 (61.5%) | 0.47 |
| Other^8^ | 115 (45.1%) | 26 (45.6%) | 74 (46.5%) | 15 (38.5%) |  |
| **Chemo*, n (%)*** |  |  |  |  |  |
| No/Unknown | 49 (19.2%) | 10 (17.5%) | 29 (18.2%) | 10 (25.6%) | 0.41 |
| Yes | 206 (80.8%) | 47 (82.5%) | 130 (81.8%) | 29 (74.4%) |  |
| **Treatment Delay*, n (%)*** |  |  |  |  |  |
| 0 | 103 (40.4%) | 23 (40.4%) | 65 (40.9%) | 15 (38.5%) | 0.96 |
| 1-6 | 96 (37.6%) | 22 (38.6%) | 59 (37.1%) | 15 (38.5%) |  |
| Other^9^ | 56 (22.0%) | 12 (21.1%) | 35 (22.0%) | 9 (23.1%) |  |

^1^ Races including Asian/Pacific Islander and American Indian/Alaska.

^2^ Sequence number of “primary only” and “1st of 2 or more primaries”, indicating aCML was the primary malignancy.

^3^ Sequence number of “2nd of 2 or more primaries”, “3rd of 3 or more primaries” and “4th of 4 or more primaries”, indicating aCML was secondary to other primary malignancies.

^4^ Marital status at diagnosis was single (never married).

^5^ Marital status of divorced, widowed and separated at diagnosis.

^6^ Median Household Income in the past 12 months (in 2021 Inflation Adjusted Dollars).

^7^ Living in a metropolitan area with a population of 1 million or more.

^8^ Including living in a metropolitan area with a population of about 250,000 to 1 million, live in a metropolitan area with a population of less than 250,000 and living in non-metropolitan areas.

^9^ Months from diagnosis to treatment is unknown or > 6.

^10^ Death attributable to aCML.

^11^ Dead of other cause such as diseases of heart, lung disease, and so on.

*HR was converted from coefficient in AFT regression model.

Abbreviations: AFT, accelerated failure time; aCML, atypical chronic myeloid leukemia; Chemo, chemotherapy; CI, confidence interval; DSS, disease-specific survival; HR, hazard ration; OS, overall survival.
