## Supplementary material for "Comprehensive Analysis of Atypical chronic myeloid leukemia (aCML): Epidemiology, Clinical Features, and Survival Outcomes Based on SEER Database Insights": Table 3

**Table 3** Univariable AFT regression analysis for OS and DSS of aCML patients

| **Parameters** | **OS** | | | **DSS** | | |
| --- | --- | --- | --- | --- | --- | --- |
|  | **HR*** | **95% CI** | ***P*-value** | **HR*** | **95% CI** | ***P*-value** |
| **Sex** |  |  |  |  |  |  |
| Male | *ref* |  |  | *ref* |  |  |
| Female | 0.99 | [0.74, 1.33] | 0.960 | 1.01 | [0.73, 1.40] | 0.940 |
| **Age** |  |  |  |  |  |  |
| <60 | *ref* |  |  | *ref* |  |  |
| 60+ | 3.59 | [2.38, 5.41] | < 0.001 | 3.17 | [2.03, 4.94] | < 0.001 |
| **Race** |  |  |  |  |  |  |
| White | *ref* |  |  | *ref* |  |  |
| African American | 0.70 | [0.43, 1.16] | 0.170 | 0.81 | [0.47, 1.39] | 0.440 |
| Other^1^ | 1.07 | [0.69, 1.68] | 0.750 | 1.33 | [0.84, 2.12] | 0.220 |
| **Marital Status** |  |  |  |  |  |  |
| Married | *ref* |  |  | *ref* |  |  |
| Single^2^ | 0.60 | [0.37, 1.00] | 0.603 | 0.57 | [0.32, 1.01] | 0.050 |
| Other^3^ | 1.13 | [0.84, 1.53] | 0.420 | 1.04 | [0.74, 1.46] | 0.840 |
| **Diagnosis Year** |  |  |  |  |  |  |
| 2001-2010 | *ref* |  |  | *ref* |  |  |
| 2011-2020 | 1.13 | [0.84, 1.53] | 0.410 | 1.08 | [0.77, 1.49] | 0.670 |
| **Sequence** |  |  |  |  |  |  |
| Primary aCML^4^ | *ref* |  |  | *ref* |  |  |
| Secondary aCML^5^ | 1.14 | [0.83, 1.57] | 0.420 | 1.11 | [0.78, 1.58] | 0.570 |
| **Income**^6^ |  |  |  |  |  |  |
| <$50,000 | *ref* |  |  | *ref* |  |  |
| $50,000-$75,000 | 0.89 | [0.57, 1.40] | 0.630 | 0.83 | [0.50, 1.36] | 0.450 |
| $75,000+ | 0.91 | [0.58, 1.44] | 0.690 | 0.91 | [0.55, 1.50] | 0.710 |
| **Residence** |  |  |  |  |  |  |
| Metropolitan > 1M^7^ | *ref* |  |  | *ref* |  |  |
| Other^8^ | 1.13 | [0.85, 1.50] | 0.390 | 1.19 | [0.87, 1.63] | 0.270 |
| **Treatment Delay** |  |  |  |  |  |  |
| 0 | *ref* |  |  | *ref* |  |  |
| 1-6 | 1.08 | [0.79, 1.49] | 0.630 | 1.05 | [0.74, 1.49] | 0.790 |
| Other^9^ | 1.05 | [0.72, 1.51] | 0.810 | 1.01 | [0.67, 1.53] | 0.960 |
| **Chemo** |  |  |  |  |  |  |
| No | *ref* |  |  | *ref* |  |  |
| Yes | 1.00 | [0.70, 1.42] | 0.990 | 1.10 | [0.74, 1.65] | 0.630 |

^1^ Races including Asian/Pacific Islander and American Indian/Alaska.

^2^ Marital status at diagnosis was single (never married).

^3^ Marital status of divorced, widowed and separated at diagnosis.

^4^ Sequence number of “primary only” and “1st of 2 or more primaries”, indicating aCML was the primary malignancy.

^5^ Sequence number of “2nd of 2 or more primaries”, “3rd of 3 or more primaries” and “4th of 4 or more primaries”, indicating aCML was secondary to other primary malignancies.

^6^ Median Household Income in the past 12 months (in 2021 Inflation Adjusted Dollars).

^7^ Living in a metropolitan area with a population of 1 million or more.

^8^ Including living in a metropolitan area with a population of about 250,000 to 1 million, live in a metropolitan area with a population of less than 250,000 and living in non-metropolitan areas.

^9^ Months from diagnosis to treatment is unknown or > 6.

*HR was converted from coefficient in AFT regression model.

Abbreviations: AFT, accelerated failure time; aCML, atypical chronic myeloid leukemia; Chemo, chemotherapy; CI, confidence interval; DSS, disease-specific survival; HR, hazard ration; OS, overall survival.
