## Supplementary material for "Comprehensive Analysis of Atypical chronic myeloid leukemia (aCML): Epidemiology, Clinical Features, and Survival Outcomes Based on SEER Database Insights": Table 4

**Table 4** Multivariable AFT regression analysis for OS and DSS of aCML patients

| **Parameters** | **OS** | | | **DSS** | | |
| --- | --- | --- | --- | --- | --- | --- |
|  | **HR*** | **95% CI** | ***P*-value** | **HR*** | **95% CI** | ***P*-value** |
| **Sex** |  |  |  |  |  |  |
| Male | *ref* |  |  | *ref* |  |  |
| Female | 1.00 | [0.73, 1.40] | 0.987 | 1.03 | [0.73, 1.40] | 0.861 |
| **Age** |  |  |  |  |  |  |
| <60 | *ref* |  |  | *ref* |  |  |
| 60+ | 3.65 | [2.34, 5.70] | < 0.001 | 3.08 | [1.92, 4.90] | < 0.001 |
| **Race** |  |  |  |  |  |  |
| White | *ref* |  |  | *ref* |  |  |
| African American | 0.89 | [0.53, 1.50] | 0.671 | 1.02 | [0.58, 1.80] | 0.937 |
| Other^1^ | 1.22 | [0.74, 2.00] | 0.438 | 1.50 | [0.89, 2.50] | 0.129 |
| **Marital Status** |  |  |  |  |  |  |
| Married | *ref* |  |  | *ref* |  |  |
| Single^2^ | 0.92 | [0.54, 1.60] | 0.765 | 0.75 | [0.41, 1.40] | 0.363 |
| Other^3^ | 1.12 | [0.81, 1.60] | 0.492 | 1.02 | [0.70, 1.50] | 0.908 |
| **Diagnosis Year** |  |  |  |  |  |  |
| 2001-2010 | *ref* |  |  | *ref* |  |  |
| 2011-2020 | 1.06 | [0.77, 1.50] | 0.711 | 1.03 | [0.73, 1.50] | 0.861 |
| **Sequence** |  |  |  |  |  |  |
| Primary aCML^4^ | *ref* |  |  | *ref* |  |  |
| Secondary aCML^5^ | 1.01 | [0.73, 1.40] | 0.938 | 1.00 | [0.69, 1.50] | 0.998 |
| **Income**^6^ |  |  |  |  |  |  |
| <$50,000 | *ref* |  |  | *ref* |  |  |
| $50,000-$75,000 | 0.84 | [0.52, 1.40] | 0.472 | 0.83 | [0.49, 1.40] | 0.487 |
| $75,000+ | 0.72 | [0.43, 1.20] | 0.214 | 0.77 | [0.44, 1.40] | 0.384 |
| **Residence** |  |  |  |  |  |  |
| Metropolitan > 1M^7^ | *ref* |  |  | *ref* |  |  |
| Other^8^ | 0.97 | [0.70, 1.30] | 0.856 | 1.07 | [0.74, 1.50] | 0.729 |
| **Treatment Delay** |  |  |  |  |  |  |
| 0 | *ref* |  |  | *ref* |  |  |
| 1-6 | 0.99 | [0.71, 1.40] | 0.941 | 0.94 | [0.65, 1.40] | 0.724 |
| Other^9^ | 0.70 | [0.38, 1.30] | 0.229 | 0.78 | [0.41, 1.50] | 0.444 |
| **Chemo** |  |  |  |  |  |  |
| No | *ref* |  |  | *ref* |  |  |
| Yes | 0.78 | [0.43, 1.40] | 0.409 | 0.95 | [0.49, 1.80] | 0.874 |

^1^ Races including Asian/Pacific Islander and American Indian/Alaska.

^2^ Marital status at diagnosis was single (never married).

^3^ Marital status of divorced, widowed and separated at diagnosis.

^4^ Sequence number of “primary only” and “1st of 2 or more primaries”, indicating aCML was the primary malignancy.

^5^ Sequence number of “2nd of 2 or more primaries”, “3rd of 3 or more primaries” and “4th of 4 or more primaries”, indicating aCML was secondary to other primary malignancies.

^6^ Median Household Income in the past 12 months (in 2021 Inflation Adjusted Dollars).

^7^ Living in a metropolitan area with a population of 1 million or more.

^8^ Including living in a metropolitan area with a population of about 250,000 to 1 million, live in a metropolitan area with a population of less than 250,000 and living in non-metropolitan areas.

^9^ Months from diagnosis to treatment is unknown or > 6.

*HR was converted from coefficient in AFT regression model.

Abbreviations: AFT, accelerated failure time; aCML, atypical chronic myeloid leukemia; Chemo, chemotherapy; CI, confidence interval; DSS, disease-specific survival; HR, hazard ration; OS, overall survival.
